# Protocol for Radiographer x AI led discharge

**DOI:** 10.1101/2025.10.10.25337734

**Authors:** Rikke Bachmann, Andreas Nexmann, Janitha Mudannayake, Janni Jensen, Sigurd S. Lauvsnes, Michael Lundemann

**Affiliations:** Radiobotics ApS, Copenhagen, Denmark; Research and Innovation Unit of Radiology, University of Southern Denmark, Odense, Denmark; Department of Radiology, Odense University Hospital, Odense, Denmark; CAI-X (Centre for Clinical Artificial Intelligence), Odense University Hospital, Odense, Denmark; Department of Radiology in Norrköping, and Department of Health, Medicine and Caring Sciences, Linköping University, Linköping, Sweden

## Abstract

**Introduction:** Emergency Department (ED) overcrowding, often exacerbated by prolonged patient length of stay (LOS), is a global challenge. Patients presenting with suspected fractures—many of whom are triaged as low-acuity, contribute significantly to the ED burden. Radiographer-led discharge (RLD), supported by artificial intelligence (AI), presents a potential strategy to streamline care, reduce LOS, and maintain diagnostic safety.

**Methods:** This multi-centre, retrospective study evaluates whether diagnostic radiographers, assisted by the radiological AI decision support tool *RBfracture 2*.*6*, can safely discharge patients with no acute skeletal or joint injury. Fifteen radiographers from three countries will independently assess 340 retrospective radiographic examinations (300 consecutive and 40 enriched with rare findings). Referral notes and AI predictions are available. Reference standards are established by consensus among three MSK radiologists/reporting radiographers. Primary outcomes include ED workload reduction (true negatives) and false negative rate. Secondary outcomes will assess AI standalone performance and inter-country comparison.

**Results:** The primary object is to evaluate whether diagnostic radiographers from three different countries, assisted by the AI tool Rbfracture 2.6, can safely reduce the emergency department (ED) workload by discharging patients without acute skeletal or joint injuries, specifically those referred with suspected bone fracture or joint dislocation.

A secondary objective is to validate the performance of RBfracture 2.6 in detecting fractures, joint dislocations, elbow effusions, knee effusions, and knee lipohemarthrosis.

## Background

Bone fractures and suspected bone fractures are among the most common reasons for patients to visit the Emergency Department (ED) ^1^. Although these injuries are often minor and radiographically evident, they typically require a full ED work up including triage, clinical examination, radiographic imaging, radiological interpretation, treatment and/or physician-led discharge. These multi-step processes can contribute significantly to Emergency Department length of stay (ED-LOS) and thereby overcrowding, even in cases without identified fracture. Stable patients with only one type of resource anticipated e.g. suspected fracture requiring x-ray imaging is the group of patients with the longest ED-LOS and causes overcrowding ^2^. Radiological investigation of diagnostic imaging is considered a major bottleneck and a cause of increased ED-LOS ^3,4^ and according to Thobaity (2025) it is recommended to streamline workflows in the ED’s that causes prolonged LOS, including radiology turnaround times ^3^. An evaluation of errors in fracture diagnosis in a Norwegian ED, during a period of two years, showed that 3.1% of all fractures were missed in the initial ED visit by the ^5^.

Overcrowding is a remaining concern in ED’s, worldwide ^6^. ED overcrowding is defined as situations where the ED function is impeded as the number of patients exceeds physical or medical staff capacity as patients are waiting to be seen, undergo assessment, undergo treatment, or waiting to be either treated or discharged ^7^. The effect of overcrowding with a high occupancy (above 90%) can cause adverse patient outcomes, treatment delays, high mortality rates and prolonged inpatient LOS ^8^. Reducing the patient’s LOS is an effective measure to reduce the risk of overcrowding in the ED ^9^.

There are multiple strategies to reduce the patient LOS in ED’s, such as introducing Triage Teams with advanced triage protocols, Fast-track pathways for minor injuries as well as rapid diagnosis ^6,8,10^. Fast-track pathways aim to accelerate care by minimizing unnecessary delays and involving specialized personnel on targeted roles and include patients with minor injuries and who do not need special care as e.g. home nursing.

An emerging strategy within such pathways is radiographer-led discharge (RLD). In this model, the radiographer, with additional training in image interpretation and reporting (reporting radiographer), assesses the images and discharges or refers the patients for treatment, without requiring physician consultation^11^. Evidence from studies suggest that RLD may reduce ED-LOS and improve patient flow while reducing patient recalls and re-attendance in the ED, due to high diagnostic accuracy among the reporting radiographers, enrolled in the strategy ^11–14^. However, there is a low low number of RLD active reporting radiographers, and to the knowledge of the authors of this study, this initiative is only activated at a few ED’s in the United Kingdom.

Concurrently, the use of artificial intelligence (AI) in medical imaging has grown rapidly. AI support tools for fracture detection have not only shown high AI stand alone performance compared to medical staff, studies have also proven how hospital staff, of different educational background and seniority, enhances their diagnostic accuracy when applied with an AI support tool ^15–17^ including Emergency Department (ED) front line staff and diagnostic radiographers ^18^.

These tools may serve as valuable decision support systems for radiographers, especially in the context of fast track ED workflows. Integrating AI assistance into radiographer-led discharge models could further enhance diagnostic confidence and streamline care. Given the convergence of ED overcrowding, the need to reduce patient LOS, the expansion of radiographer roles, and the maturation of AI technologies, a logical next step is to explore how these elements can work together. Understanding how diagnostic radiographers (not reporting radiographers), supported by AI, perform in fracture detection and patient pathway management is important for assessing the feasibility and safety of such models, before introducing this in real world settings.

### Objective

The primary objective is to evaluate whether diagnostic radiographers from three different countries, assisted by the AI tool Rbfracture 2.6, can safely reduce the emergency department (ED) workload by discharging patients without acute skeletal or joint injuries, specifically those referred with suspected bone fracture or joint dislocation.

#### Hypothesis

1. Diagnostic radiographers in conjunction with the AI tool (radiographers x AI) can discharge patients without acute findings immediately after X-ray examination, maintaining a false negative rate (missed fracture/dislocation/effusion/lipohemarthrosis) of less than 3.1%, as reported Hallas and Ellingsen (2006)^5^.
2. The AI tool alone can effectively triage patients for discharge or further ED treatment, with a comparable or improved performance in detecting the specified conditions.

#### Primary outcomes

- Reduction in the ED workload, measured by the percentage of patients safely discharged without acute findings (true negative).
- False negative rate of missed acute fractures, dislocations, effusions, and lipohemarthrosis when using the combined radiographer and AI approach.

#### Secondary outcomes

- Comparison of patient management performance across the three countries.
- Standalone performance of the AI tool in detecting fractures, dislocations, effusions, and lipohemarthrosis.
- Rate of incorrectly discharged patients when management decisions are based solely on AI findings.

#### Research questions

Can diagnostic radiographers, assisted by an AI fracture/joint dislocation detection tool, safely and effectively triage patients for discharge or ED treatment?

- Workload reduction in percentage
- Amount of patients with positive finding, incorrectly discharged
- Is there any difference in patient management performance between the three groups/countries?

What is the standalone performance of the AI tool?

- Workload reduction and amount of incorrectly discharged patients, if the patient management (discharge or return to ED) was solely based on AI findings.
- How does the AI tool perform in detection of the included findings?

### Study methods

This is a multi center study based on retrospective data, trying to resemble a true clinical workflow where diagnostic radiographers (US: *Radiologic Technologists*), assisted with an AI tool, will simulate to manage the patient workflow after radiographic examination. As the data is retrospective, no patients will be affected by the radiographers’ patient management decisions.

The study will follow the Guidelines for clinical trial protocols for interventions involving artificial intelligence: The SPIRIT AI extension^19^, The STARD-AI reporting guideline for diagnostic accuracy studies using artificial intelligence^20^ and Guidelines for Reporting Reliability and Agreement Studies^21^.

#### Data

340 radiographic examinations of the appendicular skeleton with suspicion of bone fracture or joint dislocation, excluding hip, will be included in the study. Data, which is sourced from a US based radiological image provider, is de-identified and only images, patient age, referral note, and bodypart information will be extracted. Data will be included to simulate a consecutive distribution of 300 radiographic exams presenting to an ED^22,23^. The study will also include an enriched sample of joint dislocations (10 positives), elbow joint effusions (10 positive), knee joint effusions (10 positives) and knee lipohemarthrosis (10 positives) to measure TP rate, as these findings are very rare and not measurable in a data set of 300 “consecutive” examinations. Radiographic examinations of the hip are excluded, as the data should simulate data from a ED fast-track pathway, thus immobil patients are excluded. Only radiographic examinations of patients above the age of 15 years will be included, reflecting the patient group in the fast track.

Included cases will be sampled, using a Large Language Model (LLM) to categorise cases into positive or negative for; fracture, dislocation, elbow effusion, knee effusion, and knee lipohemarthrosis for each of the included body parts. Afterwards, the first author of this study with a background as a reporting radiographer, will curate the dataset based on the inclusion and exclusion criteria as set below.

#### Considerations on sample sizes

The sample size of 300 (+40 enriched) included radiographic examinations is feasible from a reader point of view and based on published reader studies with an AI support tool as intervention ^18,24^. Including 5 radiographers from each of 3 countries, and assuming a true mean False Negative Rate of 2.8% with a variation of ±0.1between raters would achieve a power > 80% with a significance level of 5% to prove that diagnostic radiographers x AI perform significantly lower than 3.1%.

**Table 1:** Inclusion and exclusion criteria.

| Inclusion | Exclusion |
| --- | --- |
| • Patients $\geq 15$ years of age, referred to x-ray imaging from the Emergency department with a suspected acute bone fracture or joint dislocation injury in the appendicular skeleton, including all types of (potential) fractures e.g. traumatic, stress, periprosthetic, and pathological<br>• Radiographic examinations of the hand, wrist, forearm, elbow, humerus, shoulder, clavicle, knee, lower leg, ankle, and foot were included. Numbers of included cases aimed to resemble a normal distribution of body parts radiographed with suspected bone or joint injury in a typical emergency department<sup>24,25</sup> (See appendix 1 for the aim of included examinations)<br>• Modality is digitally acquired radiographs (Computed Radiography (CR) or Digital Radiography (DR/DX)) | • Examinations including spine, skull, face, pelvis or hip<br>• Radiographs from follow-up patient examinations<br>• Radiographic examinations that only contain one image<br>• Any patient whose radiographs were previously included in the training, tuning, or verification in the development of RBfracture 2.6.0<br>• Any patient whose radiographs were previously included in a reader study conducted by Radiobotics<br>• Studies including multiple body parts (e.g. knee and hand) |

#### Reference Standard

The Reference Standard will be established in the Collective Minds Radiology Image platform by three certified Radiologists or Reporting Radiographers, one from a Swedish hospital, one from a Dutch hospital, and one from a Danish hospital. In cases of discrepancies, where all three radiologists do not agree in presence of pathologies and/or patient management, a meeting between the three radiologists will be arranged for them to reach consensus (Reference Standard). If a consensus cannot be reached, the case will be excluded in the analysis.

Referral notes will be available for all cases. Original radiological reports and AI outputs will NOT be available to the radiologists involved in setting the Reference Standard. The pathology annotation is to assess the AI stand alone performance and to get an estimate on how many patients, without fracture, dislocation, effusion, and/or lipohemarthrosis, that will be returned to ED for further evaluation or treatment. In such cases, the radiologist will be asked to state the reason for returning the patient to the ED in the annotation. The Reference Standard will not state how many of the same pathological findings are present in one examination or the localisation, only if the exam is positive or negative for each finding.

Possible Reference Standard (RS) in test set

1. **Discharge**
  1.a. **Reference Standard negative** → No acute/subacute fractures, joint dislocation, elbow joint effusion, knee joint effusion, knee lipohemarthrosis annotated. These exams will be labeled “No fracture/dislocation/effusion/lipohemarthrosis”, to ensure the researcher did assess the radiographic exam, and not skipped it by accident.
  1.b. **Reference Standard positive** → A minimum of one acute/subacute fracture, joint dislocation, elbow joint effusion, knee joint effusion, knee lipohemarthrosis is annotated with an injury type label one for each positive finding.
2. **Return to ED**
  2.a. **Reference Standard negative** → No acute/subacute fractures, joint dislocation, elbow joint effusion, knee joint effusion, knee lipohemarthrosis annotated. These exams will be labeled “No fracture/dislocation/effusion/lipohemarthrosis”, to ensure the researcher did assess the radiographic exam, and not skipped it by accident.
  2.b. **Reference Standard positive** → A minimum of one acute/subacute fracture, joint dislocation, elbow joint effusion, knee joint effusion, knee lipohemarthrosis is annotated with an injury type label one for each positive finding.

*Note: If the reference standard radiologist chooses the “Patient to ED” tag in an exam also tagged with “No fracture/dislocation/effusion/lipohemarthrosis”, the radiologist must include a free text in the management tag, stating the reason for returning the patient to the ED*.

*Note: Patients with acute knee trauma and patients with suspected finger injuries must always be Returned to ED, to reflect normal clinical practice*.

#### Radiographer x AI readings

5 diagnostic radiographers, with a minimum of two years of experience working with Direct Radiography (DR/DX) in injury patients from the ED, will be appointed by each of the three hospital sites.

The 15 diagnostic radiographers will get access to Collective Minds Radiology Image platform, containing the curated data set of 340 radiographic examinations, including AI outputs for decision support. The radiographers will not receive face to face training on the use of the AI tool, but will receive written guidance on how to interpret the AI outputs and how to operate the Radiology platform. They will also be asked to sign an Informed Participant Sheet, explaining the study, before study initiation. For each radiographic examination, the radiographer must decide whether to discharge the patient or return the patient to the ED for further assessment/treatment. Even though the radiographer is assisted with the AI tool, it is up to the radiographer how to manage the patient - ED/discharge.

*Note: Patients with acute knee trauma and patients with suspected finger injuries must always be Returned to ED, to reflect normal clinical practice*.

The radiographer will have the referral notes, original images, and the AI predictions available in all 340 cases.The original radiology report and the Reference Standard will NOT be available to the radiographers.

Possible radiographer x AI annotations:

1. **Discharge patient (Negative)**
2. **Return patient to ED (Positive)**

#### Physical study setting

To simulate normal working conditions, reference standard annotations and radiographer annotations will be performed during normal working hours. Also, reference standard annotations must be performed using diagnostic monitors, whereas patient management annotations by radiographers can be done in any available screen in the included radiology departments, with a minimum size of 27 inches. Radiographer time spent on patient management decisions, will be recorded in the annotation platform. Recorded time will not cover the time spent on patient communication, but it can give insights on the radiographer time burden.

#### AI tool

The AI tool used in the study (RBfracture 2.6.0™ Radiobotics, Copenhagen, Denmark) is CE-marked as a class IIa medical device and is intended to be used in a clinical setting as a support tool for detecting fractures in the appendicular skeleton and knee and elbow effusions on patients ≥ 2 years of age, as well as knee lipohemarthrosis and joint dislocation (not dislocated joint replacements) in the appendicular skeleton on patients above the ≥ 15 years of ages. The AI tool is supplied with a series of radiographs as input and as output creates a Summary report and secondary captures where the detected findings are highlighted with a bounding box and finding in text as well as the confidence score, e.g. fracture 96%. Low confidence findings of 75-94% are marked with a dashed line bounding box and high confidence findings 95-99% are marked with a solid line bounding box.

The AI tool is based on a data set of more than 350,000 radiographs, collected from more than 1000 radiology departments across multiple continents. Prior to development, the data set was split into 3 subsets, a training subset consisting of 80%, a tuning subset of 10%, and an internal verification set consisting of 10% of the data.

Possible AI outputs:

1. **AI negative** → No acute/subacute fractures, joint dislocation, elbow joint effusion, knee joint effusion, knee lipohemarthrosis. A summary report with a grey dot or a grey circle. Patient management - **Discharge patient**
2. **AI positive** → A minimum of one acute/subacute fracture, joint dislocation, elbow joint effusion, knee joint effusion, knee lipohemarthrosis is detected. These outputs will have a summary report with a red dot and image overlays containing box(es) with AI confidence bounding the finding(s) and an analysis box highlighting the findings with text. Patient management - **Return to ED**

**Figure 1.**
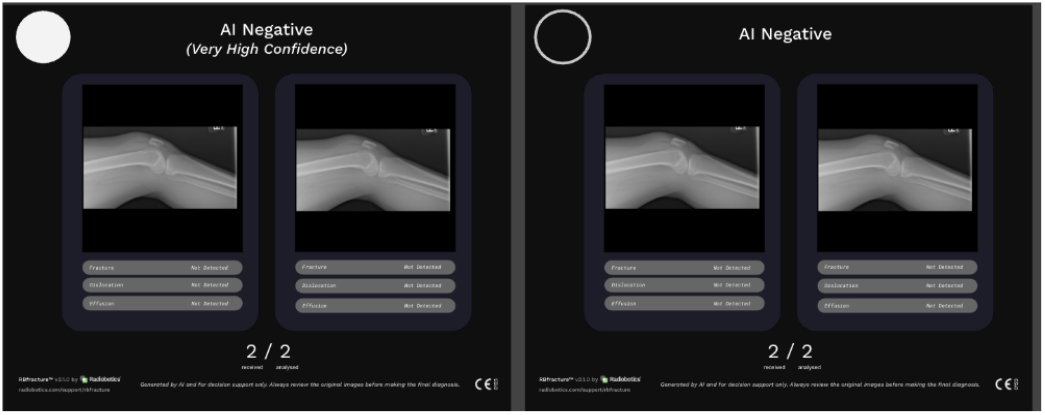
Returned summary report without findings, indicated with a grey dot or a grey circle, depending on the model confidence. In case of no findings, no images but only the summary report will be returned to the user.

**Figure 2.**
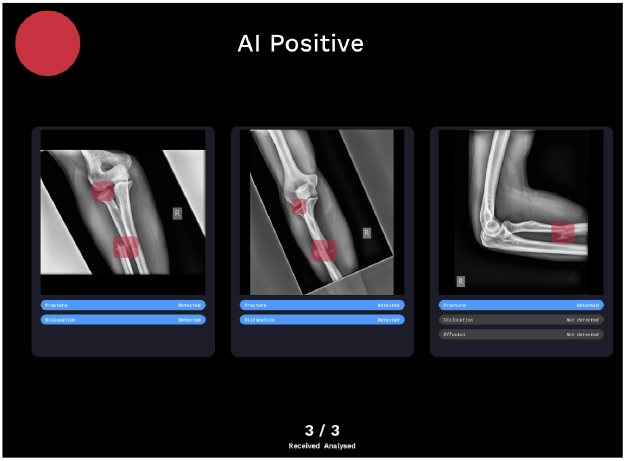
Summary report returned with positive findings. The summary report also shows how many images were sent and analysed.

**Figure 3.**
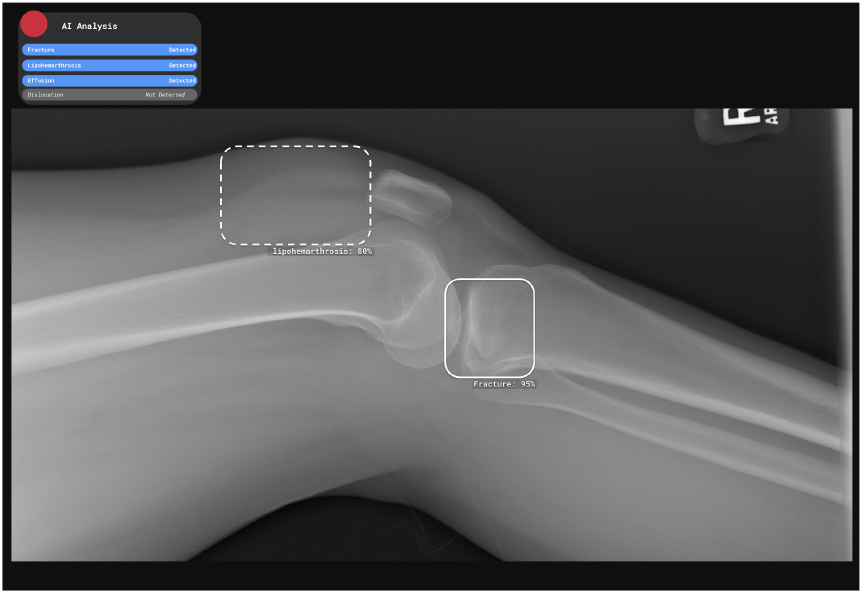
Examples of a positive case including Red dot, bounding boxes with confidence scores and text in the analysis box. Note that confidence predictions of 75-94% are marked with a dashed line box and confidence predictions of 95-100% are marked with a solid line box.

### Statistical analyses

#### Performance endpoints

Endpoints will be calculated using the Reference Standard as a benchmark for:

- AI standalone, patient management (consecutive 300 cases) and injury detection (enriched 340 cases)
- Each Radiographer x AI, patient management (consecutive 300 cases)
- Average Radiographer x AI, patient management (consecutive 300 cases)
- Average Radiographer x AI per hospital site, patient management (consecutive 300 cases)

#### Patient Management Classification

Each of the 340 cases from both the Radiographer x AI and AI standalone groups will be classified based on the Reference Standard into:

**Table 2:** Patient Management Classification metrics.

|  |  |
| --- | --- |
| True Positive (TP) | Patients correctly returned to the ED / AI Positive |
| True Negative (TN) | Patients correctly discharged / AI Negative |
| False Positive (FP) | Patients incorrectly returned to ED / AI Positive |
| False Negative (FN) | Patients incorrectly discharged / AI Negative |

The TN and FN counts will indicate the immediate workload reduction, while FN cases will undergo a quality evaluation to assess potential clinical consequences of missed injuries. Percentage wide spread of AI negative and AI negative -very high confidence in FN and TN categories for both AI stand alone and Radiographer x AI will be calculated to identify possible advantages or disadvantages for the negative classification feature.

Patient management in False Negative Rate (FNR), Proportion Discharged (PD), and Negative Predictive Value (NPV) will be calculated on the consecutive data set (300 cases) as NPV is prevalence-dependent and to reflect the normal workflow. This will be calculated for each Radiographer x AI, average Radiographer x AI per Hospital site, overall average Radiographer x AI, as well as AI standalone and compared across groups.

Radiographer x AI performance in the 40 cases with rare findings, will be discussed but not accounted for in the results, as these do not reflect the normal consecutive ED visits. Nevertheless, it is still interesting to investigate how radiographers perform on such rare cases.

#### Comparative Analyses

Performance differences will be assessed between:

- AI standalone vs. Average Radiographer x AI, patient management (consecutive 300 cases)
- Average Radiographer x AI across the three hospital groups (consecutive 300 cases)

Comparative analyses will be conducted using statistical tests such as chi-square tests for categorical data and t-tests or ANOVA for continuous data, where appropriate. Adjustments for multiple comparisons will be made using the Bonferroni correction.

#### Injury detection, AI standalone

All 340 examinations will undergo a naive injury wise performance evaluation of the AI support tool. All possible findings in each examination will be classified as True Positive (TP), True Negative (TN), False Positive (FP), and False Negative (FN). Effusions will only be assessed and classified in lateral images of the elbow and knee and lipohemarthrosis only on lateral knee views. This means that one case, e.g. a wrist can be classified as True Positive for a fracture finding, False Negative for a missed dislocation, and not be assessed for effusion and lipohemarthrosis, thus the results will show more T/F classifications, than number of included exams.

**Table 3:** Injury detection classification metrics. RS - Reference Standard.

|  |  |
| --- | --- |
| True Positive (TP) | An AI prediction also deemed positive for the same injury by the RS |
| True Negative (TN) | Each non AI predicted injuries in each exam where the injury is not stated by the R |
| False Positive (FP) | An AI predicted injury in an exam where the same injury is not stated by the R S |
| False Negative (FN) | A RS stated injury not predicted by the AI tool |

#### Quantifying Uncertainty

Uncertainty in estimates will be quantified using 95% confidence intervals (CIs) for all key metrics, providing a range within which the true value is expected to lie with 95% confidence.

#### Visualization

The average Radiographer x AI performance on the consecutive data set, including TP, TN, FP, FN (with missed findings), will be visualized using appropriate figures

#### Performance Table

Performance in patient management for each radiographer x AI, average Radiographer x AI per included Hospital and overall Radiographer x AI average as well as AI standalone performance in False Negative Rate (FNR), Proportion Discharged (PD), and Negative Predictive Value (NPV) will be presented in a combined table including the 300 consecutive data set.

AI standalone performance in pathology detection will be presented in a table including performance metrics per injury and in total, including the 340 enriched data set.

#### Subgroup analysis

Age will be reported per patient to investigate if there are any differences in performance for different age groups. Age will be reported as median and range, sex (male/female) and symptomatic side (left/right) and body part are the only patient-related variables that are extracted from the PACS system. These variables will be presented in general and not on an individual patient level, thus identification of individual patients is not possible.

## Ethics and Dissemination policy

Ethical approval for this study was obtained from “Danish Research Ethics Committees”, case number: 2501870. The Institutional Review Board WCG Clinical Service determined that the study is considered exempt from full IRB approval for patient consent because the included data is fully anonymised on 2025.07.15, IRB tracking number: 20252523.

This study will be conducted in accordance with the Declaration of Helsinki. The General Data Protection Regulation and the Danish Data Protection Act will be respected and the data is in compliance with HIPAA. The study protocol follows the SPIRIT-AI checklist ^19^. Collaboration agreements are established between Radiobotics and the data provider and will be established between Radiobotics and each of the three Hospitals sites before study initiating.

The study did not receive any funding and there were no financial transactions between the parties involved in this project.

The authors RB, JM, AN, and ML are all employees at Radiobotics, the manufacturer of RBfracture, the AI decision support tool used in the study.

Findings from this study are intended for publication in scientific peer-reviewed journals. Furthermore, results will be presented at national and international conferences with oral and/or poster presentations. The first author assumes responsibility for all practical issues and first drafts of articles. The article is written and authorship decided according to guidelines by the International Committee of Medical Journal Editors.

## Data Availability

All data produced in the present study are available upon reasonable request to the authors

## Appendix 1

*Gray column: These are positives and also included in fracture, dislocation, fracture+dislocation and is therefore not included in the total number of positives. Effusion/FBI - fracture/dislocation should be considered as positive (return patient to the ED) and the image does not include visible fracture or dislocation*.

Appendix 1
| Desired Consecutive data (simulating) |  |  |  |  |  |  |  |
| --- | --- | --- | --- | --- | --- | --- | --- |
| Study-type | Fracture | Dislocation | Fracture and dislocation | Effusion/Lipo (+frac/disl) | Effusion/Lipo (-frac/disl) | Negative | Total (%) |
| Hand | 15 | 2 | 2 |  |  | 43 | 62 (20%) |
| Wrist | 12 |  | 1 |  |  | 27 | 40 (14%) |
| Forearm | 0 |  |  |  |  | 2 | 2 (1%) |
| Elbow | 5 | 1 | 1 | 4 | 3 | 20 | 30 (10%) |
| Humerus | 1 |  |  |  |  | 3 | 4 (1.3%) |
| Shoulder | 3 | 1 | 1 |  |  | 23 | 28 (9%) |
| Knee | 2 | 1 | 0 | 2 | 1 | 20 | 24 (8%) |
| Lower leg | 1 |  |  |  |  | 9 | 10 (3.3%) |
| Ankle | 8 |  | 2 |  |  | 40 | 50 (17%) |
| Foot | 10 | 0 | 1 |  |  | 39 | 50 (16%) |
| Total | 57 | 5 | 8 | 6 | 4 | 226 (75%) | 300 |

*Number of enriched exams with positive findings as listed. Numbers in parentheses are the total number of exams with positive findings in the consecutive and the enriched data set*.

| Desired Enriched data |  |  |  |
| --- | --- | --- | --- |
| Study-type | Joint effusion<br>(Incl. cons.) | Lipohemarthrosis<br>(Incl. cons.) | Joint dislocation<br>(Incl. cons.) |
| Hand |  |  | 2 (6) |
| Wrist |  |  | 0 (1) |
| Elbow | 10 (17) Some will<br>include fracture as<br>well |  | 2 (4) |
| Shoulder |  |  | 2 (4) |
| Knee | 10 (12) | 10 (11) | 2 (3) |
| Ankle |  |  | 1 (3) |
| Foot |  |  | 1 (2) |
| Total | 20 (29) | 10 (11) | 10 (23) |

